# Impact of a new SARS-CoV-2 variant on the population: A mathematical modeling approach

**DOI:** 10.1101/2021.02.24.21252406

**Authors:** Gilberto Gonzalez-Parra, David Martínez-Rodríguez, Rafael-J. Villanueva-Micó

## Abstract

**S**everal SARS-CoV-2 variants have emerged around the world and the appearance of other variants depends on many factors. These new variants might have different characteristics that can affect the transmissibility and death rate. The administration of vaccines against the coronavirus disease 2019 (COVID-19) started in early December of 2020 and in some countries the vaccines will not soon be widely available. In this article, we study the impact of a new more transmissible SARS-CoV-2 strain on prevalence, hospitalizations, and deaths related to the SARS-CoV-2 virus. We study different scenarios regarding the transmissibility in order to provide a scientific support for public health policies and bring awareness of potential future situations related to the COVID-19 pandemic. We construct a compartmental mathematical model based on differential equations to study these different scenarios. In this way, we are able to understand how a new, more infectious strain of the virus can impact the dynamics of the COVID-19 pandemic. We study several metrics related to the possible outcomes of the COVID-19 pandemic in order to assess the impact of a higher transmissibility of a new SARS-CoV-2 strain on these metrics. We found that, even if the new variant has the same death rate, its high transmissibility can increase the number of infected people, those hospitalized, and deaths. The simulation results show that health institutions need to focus on increasing non-pharmaceutical interventions and the pace of vaccine inoculation since a new variant with higher transmissibility as, for example, VOC-202012/01 of lineage B.1.1.7, may cause more devastating outcomes in the population.

## 1 Introduction

The world is currently suffering one of the worst pandemics in history. The spread of the SARS-CoV-2 virus started in Wuhan, China and has affected now the whole world in one way or another. Recently, several variants of the SARS-CoV-2 virus have been detected. These variants are defined depending on the number and type of mutations [66, 75]. There are many concerns about what the characteristics of these new variants are regarding infectiousness and severity of disease. For instance, the public, researchers, and media are asking what the consequences are of having a more transmissible SARS-CoV-2 variant [70, 73, 90, 34]. During the last month of 2020 a few countries started with the inoculation of vaccines against the SARS-CoV-2 virus, but it seems that for many them the vaccines will not be available for at least the first semester of 2020 [35, 127, 78, 117, 99, 129]. This pandemic has caused more than 100 million confirmed cases and more than 2.2 million deaths [11, 59]. The SARS-CoV-2 virus is not the most transmissible virus, but it is very dangerous since it causes in some cases severe pneumonia and death [80, 98]. The spread of the SARS-CoV-2 virus is influenced by many factors that are currently not fully well understood [81, 110, 138, 98, 22, 23]. Among these, it is known that social behavior, mobility, age, weather variables, and virus mutation can affect the dynamics of the transmission of the SARS-CoV-2 virus in the human population [152, 108, 147]. As well, there may be further unknown factors that may affect the spread of the SARS-CoV-2 virus. The mutations of viruses are common and, as a consequence, the SARS-CoV-2 can acquire mutations with fitness advantages and immunological resistance [66, 55]. Several researchers have devoted time to study evolutionary transitions in order to ensure effectiveness of the vaccines and immunotherapeutic interventions [151, 43, 100, 157, 66]. A new variant of the SARS-CoV-2 virus has been detected in England, and is the VOC-202012/01 of lineage B.1.1.7. [106, 76, 33, 34]. VOC-202012/01 is characterized by multiple mutations in the spike protein. Among them, N501Y is of major concern because it involves one of the six key amino acid residues determining a tight interaction of the SARS-CoV-2 receptor-binding domain (RBD) with its cellular receptor angiotensin-converting enzyme 2 (ACE2) [33, 149]. It has been found by phylogeny that viral strains obtained from a patient belonged to the B.1.1 lineage. The authors in [33] used a time-scaled maximum likelihood tree that suggests that spike N501T variants emerged in early August in northern Italy.

From October 2020 the number of infected cases and deaths increased dramatically in England. It has been found that the new SARS-CoV-2 variant VOC-202012/01 was prevalent and its proportion increased during the recent months in England [103, 55]. It is not clear if a single or a combination of different mutations would change the viral transmissibility, virulence, clinical and epidemiological presentations, or vaccine efficacy [106]. However, several researchers and institutions have mentioned that the new SARS-CoV-2 variant VOC-202012/01 is more transmissible than the previously prevalent variants [76, 111, 139, 34, 55]. In particular, in [76] the authors found that variant VOC-202012/01 is estimated to present an ℛ, 1.75 times higher than 501N, meaning it is 75% more transmissible compared with the 501N strain. Furthermore, this variant VOC-202012/01 has become the dominant strain in England in November/December 2020. It is expected that additional mutations will appear around the world and possibly even more after global vaccination due to mutation pressure [106]. Therefore, studying the impact of new strains of the SARS-CoV-2 virus is of paramount importance.

Mathematical models have been useful to study and understand the dynamics of many infectious diseases. These models, in conjunction with statistical analyses and computational techniques, are valuable tools to test hypotheses and analyze the impact of factors on the infectious disease processes. *In silico* simulations of the mathematical models allow the investigation of different potential scenarios related to particular epidemics [89, 8, 112, 38, 3, 113, 74, 135, 62, 61]. The outcomes of the complex infectious disease processes under different scenarios are generally impossible to predict without mathematical models and computational techniques. Even with these tools, there are some caveats and weaknesses, due to several reasons, including for instance implicit hypotheses and/or unreliable data [115, 48, 57, 124]. The results of the numerical simulations might be counter-intuitive and thus interesting from a point of view of deeper understanding. For instance, it has been found that people have difficulty understanding a simple process of balance of energy[1]. This last study showed that people exhibit systematic errors indicative of the use of erroneous heuristics [1]. Thus, mathematical models help to provide knowledge and scientific support to make the best decision from a public health point of view. In this study, we provide a mathematical model to study the impact that a more transmissible SARS-CoV-2 variant can have on health metrics such as prevalence, hospitalization, and deaths.

Mathematical modeling in combination with computational and statistical techniques have been used to study the spread of the SARS-CoV-2 virus [68, 125, 32, 22, 130, 69, 155]. Several of these models are on SIR-type (Susceptible-Infected-Recovered) based[101, 5, 107, 12]. However, other researchers have used susceptible-exposed-infected-recovered (SEIR) type models [42, 130, 50, 153, 83]. One advantage of mathematical models is that many different simulations can be performed and this allows us to study the dynamics of a disease under a variety of complex scenarios. We and other researchers have found that many forecasts related to the COVID-19 pandemic disagree with each other due to many uncertainties in key characteristics related to the spread of the SARS-CoV-2 virus in the human population [115, 48, 30, 57, 124, 134, 68, 125, 32, 130, 69, 155]. Currently we are facing new variants of the SARS-CoV-2 virus. This has raised questions and increases the uncertainty in the dynamics. It has been found that the SARS-CoV-2 is mutating and its transmission is more efficient [151, 94, 24, 77]. There is a growing literature about new variants of the SARS-CoV-2, but it is, of course, not clear what further variants will occur in the future [151, 43, 100, 157, 66]. For instance, using analysis of the frequency of the 614D and 614G variants over time it has been suggested that locations that reported 614D viruses early in the pandemic were often later dominated by 614G viruses [66].

In this study, we construct a compartmental mathematical model of SARS-CoV-2 transmission and disease progression. We use *in silico* simulations to study different scenarios related to SARS-CoV-2 transmissibility and disease severity, considering two variants. In particular, we assume that the second variant is the VOC-202012/01 of lineage B.1.1.7. However, the mathematical model can be used for any other new variant. We include the asymptomatic carriers of the SARS-CoV-2, who are nevertheless able to spread the virus. The reason to include them is that asymptomatic people are very important contributors in the spread of the SARS-CoV-2 [4, 51, 87, 96, 122, 36, 63, 44, 132, 22]. It has been found that quantitative SARS-CoV-2 viral loads were similarly high for infected individuals with symptoms, pre-symptomatic, or asymptomatic. In addition, 6 to 24 times more estimated infections per site have been found with seroprevalence tests than the number of reported cases with coronavirus disease 2019 (COVID-19) [46]. In this study we explore different potential scenarios with different transmissibilities of the SARS-CoV-2.

This paper is organized as follows. In Section 2, we present the mathematical model of SARS-CoV-2 transmission and disease progression. In Section 3, the numerical simulation results using the constructed mathematical model of SARS-CoV-2 transmission and disease progression are shown, and the last section is devoted to discussion and conclusions.

## 2 Materials and Methods

### 2.1 Mathematical Model

As has been mentioned in the introduction, providing quantitative forecasts for COVID-19 pandemic has been very challenging and some criticism has arisen [115, 48, 30, 57, 124, 134, 68, 125, 130, 69, 155]. In this study, we focus on the qualitative impact that a new more transmissible SARS-CoV-2 variant could have on the population. Using the mathematical model and numerical simulations we can study the potential consequences of the more highly transmissible variant. A very complex mathematical model that represents more closely the real world situation can be constructed, but the number of parameters of such model would be very large and moreover the values of some of these parameters would increase the overall uncertainty. We propose a mathematical model that approximates reality but with some limitations in order to produce scientific results that bring awareness to health institutions and people. Moreover, we provide answers brought **bt** (what is this??) media and people regarding the impact of a new highly transmissible SARS-CoV-2 variant.

We construct a compartmental model based on differential equations that considers two variants of the SARS-CoV-2 virus. In particular we assume that the second variant is the VOC-202012/01 of lineage B.1.1.7. The model includes individuals in the susceptible, latent, infected, asymptomatic, and hospitalized stages. The transitions of individuals between the stages depend on the COVID-19 progression. The model also considers that individuals only can get one SARS-CoV-2 variant. Thus, we can classify individuals in two disjoint groups related to disease progression: those infected with variant 1 and those infected with variant 2. The model considers that individuals once infected with a SARS-CoV-2 variant have full cross-immunity against the other variant due to the adaptive immune response. The model has individuals in the latent stage (either with previous variant 1 or new variant 2) who are not yet infectious. The individuals remain in the latent stage for a certain time with mean *α*. The individuals then transit into the infective symptomatic or asymptomatic stages (either with variant 1 or variant 2), where they are able to spread the SARS-CoV-2 virus to other individuals. They stay in the infectious stage for a certain time with mean *γ*. After that, individuals in the asymptomatic stage move to the recovered stage. However, individuals in the infective symptomatic stage can move to the recovered or to the hospitalized compartments, depending on the level of disease progression. This classic type of model assumes implicitly exponential distributions, but the Erlang distributions are usually more realistic at the expense of more complexity in the models and more parameters [56, 41, 114, 40, 30, 141]. Thus, many studies use models based on differential equations that assume exponential distributions to avoid greater complexity in the models and in the analysis. However, in many cases exponential distributions are not far from reality. We have found that the length of stay in the hospital is not far from an exponential distribution [29]. Finally, the model considers that hospitalized individuals may die due the COVID-19 disease [29, 31, 32, 156]. This last metric (or outcome) is of paramount importance for health institutions and people since it is irreversible [32, 137, 142, 150]. Mathematical models have proven valuable in understanding many infectious diseases. However, it is important to mention that any model-based analysis has methodological limitations. The type of model presented here assumes implicitly homogeneous mixing in the population [89, 8]. The homogeneous mixing has been assumed for other viruses less transmissible than SARS-CoV-2 like HIV or influenza, predictions should be taken cautiously. Despite some classical limitations this type of mathematical model provides ways to understand the qualitative dynamics of some infectious diseases under certain conditions. In this way, mathematical modeling allows us to gain valuable insight into a disease and its transmission.

We use a mathematical model similar to a *SEIR*-type epidemiological model to understand the dynamics of the SARS-CoV-2 virus spread on the human population under two variants of the SARS-CoV-2 virus. The mathematical model and general methodology can be extrapolated to more variants increasing the number of differential equations and parameters. The constructed model has parameters that can be varied in order to study different potential scenarios in order to take into account the uncertainty of some processes of the disease progression. For instance, the transmissibility of the variants of SARS-CoV-2 virus can be modified. This is important since recently it has been mentioned that new variants might have significant differences in transmissibility [66, 34, 90]. In addition, the models consider parameters related to the severity of the disease, which includes the hospitalization rate for the different strains. However, currently it is not known if the new variants have different mortality and/or hospitalization rates. Finally, we develop this mathematical model to study the impact of one new SARS-CoV-2 variant on prevalence, hospitalizations, and deaths related to the SARS-CoV-2 virus. It is important to remark that the mathematical model does not consider in-person mutation so that the person may be able to transmit a different SARS-CoV-2 variant.

The constructed mathematical model based on differential equations is given by

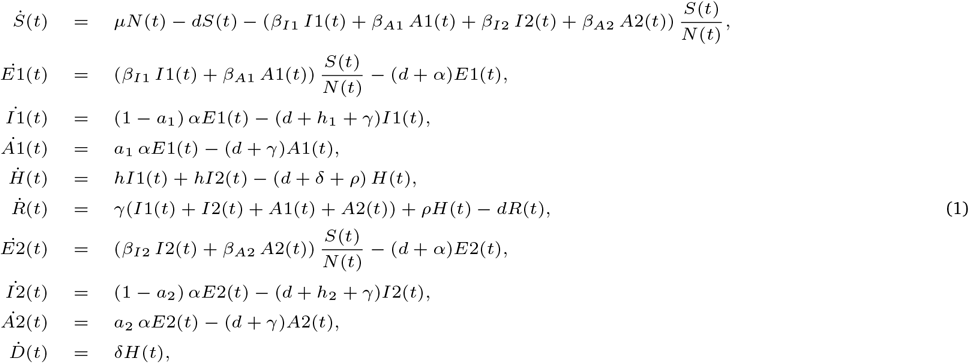

where *Ei*(*t*), *Ii*(*t*) and *Ai*(*t*), denote the latent, infective symptomatic and asymptomatic for individuals infected with variation *i* respectively. The other variables *S*(*t*), *H*(*t*), *R*(*t*) and *D*(*t*) represent susceptible, hospitalized, recovered and deaths at time *t*. The mathematical model assumes that people in states *Ei*(*t*), *H*(*t*), and *R*(*t*) do not transmit the infection. For instance, we assume that health care workers and other people in contact with hospitalized individuals follow strict guidelines. The constructed model considers that COVID-19 confers full cross-immunity after recovery. The Figure 1 depicts the flow of individuals from one subpopulation to another.

**Figure 1:**
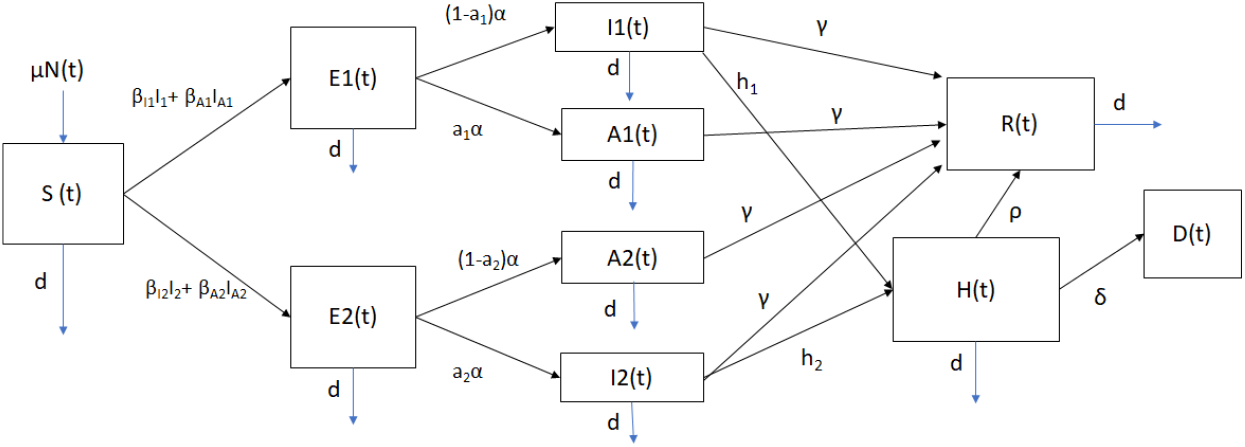
Diagram for the COVID-19 mathematical model (1). The boxes represent the subpopulation and the arrows the transition between the subpopulations. Arrows are labeled by their corresponding model parameters.

### 2.2 Parameter values

In this study we are interested in understanding the qualitative impact of a more transmissible new SARS-CoV-2 variant on the dynamics of the COVID-19 pandemic. In particular, we consider that the second variant is the VOC-202012/01 of lineage B.1.1.7. However, the general methodology and mathematical model could be applied to other SARS-CoV-2 variants. Taking more accurate parameter values would provide more robust qualitative results, even though small differences would not change the qualitative conclusions. We assume that the rates of virus transmission in asymptomatic and symptomatic individuals are constant from the beginning of the simulation. This assumption implies that people would keep approximately the same behavior (on average) regarding SARS-CoV-2 virus spread protection. This assumption is reasonable but may change if the number of cases and deaths increase dramatically due to the new SARS-CoV-2 variant. It is important to mention that in countries where a vaccination program is advancing quickly it is necessary to construct another model that considers the vaccinated population [95, 27, 53, 28, 82]. In addition, if the behavior of the people changes it would be more realistic to include time-varying transmissibility, which has been used to study other infectious diseases [131, 72, 68, 62, 28]. Doing this would make the mathematical modeling process more complex since the estimation of a time-varying parameter is more difficult to achieve, and identifiability issues might arise. Furthermore, in this study we cannot predict how the behavior of individuals might change in the future. Thus, we take an approximation and the conservative assumption that the transmissibility of the two SARS-CoV-2 variants would not change over the simulation which seems plausible [95]. However, the number of effective contacts between the susceptible and infected vary over time due to the variation of these subpopulations (for more details see [8, 47, 26, 25]).

We assume that the parameters related to the latent and infectious stages are the same for both SARS-CoV-2 variants. Some studies have indicated that the antibody titers may decline over time in patients recovered from COVID-19, particularly in those who were asymptomatic [144]. However, we do not consider that recovered individuals can return to the susceptible stage. Further studies are needed to know the immunity period. In addition, we consider a short time period (1 year) since we expect that after one year all the countries would have a fully developed vaccination program, and another model that includes vaccination would be necessary [82, 53, 95, 120].

For the death rate of hospitalized individuals we use data from scientific sources [32, 60, 148, 88, 95]. We used the weighted average of the probability of dying for severe and critical cases (ICU), and in addition we took into account the average length of stay in the hospital [95]. We varied in a reasonable way the death rate in order to take into account the possible uncertainty in the data. As we have mentioned above the qualitative results would not change with small differences in the death rate and hospitalization rates. Even so, we try to assume parameter values as accurate as possible.

For the asymptomatic cases and proportions we also relied on data from the scientific literature [11, 13, 84, 32, 63, 87, 92, 91, 154]. However, the discrepancies in the relevant data are great. We chose as a conservative starting point that the percentage of infections that are asymptomatic is 50% [11]. However, for the numerical simulations we additionally considered a range around 50% [11, 91]. Although the results of this study can be extrapolated to several different countries we specifically chose Colombia since it has a mid-size population, a full vaccination program that has not started yet, and it has a significant number of infected cases. It is important to remark that the only parameters that can be considered specific for Colombia are the birth and death rates. Thus, the approach can be used for other countries if these parameters are changed. Another important point is that these parameters related to demographics have a very small impact on the results since the time scale of the demography rates are much slower than the disease [47]. The values of the parameters for the base scenario are given in Table 1.

**Table 1:** Mean values of parameters used to perform numerical simulations of the different scenarios.

| Parameter | Symbol | Value or range |
| --- | --- | --- |
| Latent period | $\alpha^{-1}$ | 5.2 days [78, 71, 131] |
| Infectious period | $\gamma^{-1}$ | 7 days [78] |
| Hospitalization rate | $h^{-1}$ | $3.5 \text{ days} \times 0.04^{-1}$ [32, 78, 49] |
| Hospitalization period | $\rho^{-1}$ | $10.4 \times 0.897^{-1} \text{ days}$ [32, 78, 49] |
| Death rate (hospitalized) | $\delta^{-1}$ | $10.4 \text{ days} \times 0.103^{-1}$ [104, 95] |
| Probability of being asymptomatic | $a$ | [0.2 – 0.8] [11, 91] |
| Birth rate | $\mu^{-1}$ | 0.0000407726 days [133] |
| Death rate | $d^{-1}$ | 0.00001523835 days [133] |

In every country in the world there are several non-pharmaceutical interventions and people are aware of the current COVID-19 pandemic. We assume for the numerical simulations that the new SARS-CoV-2 variant is more transmissible. Thus, we consider that *β*_*I*1_ *< β*_*I*2_, and we take *β*_*I*2_ = *pβ*_*I*1_ where *p >* 1. We vary the value of *p* to include the uncertainty in the transmissibility of the new SARS-CoV-2 variant, and analyze the impact on some crucial outcomes. Regarding asymptomatic carriers we follow the same trend assuming that *β*_*A*1_ *< β*_*A*2_, and *β*_*A*2_ = *pβ*_*A*1_ where *p >* 1. For the infectiousness of the asymptomatic carriers we vary the transmission rate parameter *β*_*A*1_, but taking into account that it is similar or smaller than the infectiousness of symptomatic carriers [13, 84, 32, 63, 118, 154]. One article surprisingly found that asymptomatic carriers have a higher viral load, and, that could mean that asymptomatic carriers might have larger infectiousness than symptomatic people [45]. We disregard this scenario due to no further evidence of this situation. For the *β*_*s*_ parameters we assume values in the range of [0.1− 1.5], which are values found in different studies, even though certainly it has uncertainty and variation for each region or country [95, 53, 73, 79, 128, 28].

### 2.3 Initial conditions for the scenarios

As has been mentioned before, for the initial conditions we assume the initial population of Colombia which is relatively close to the population of several countries that are not expected to have a full vaccination program soon. We rely on data from the scientific literature and demographics from Colombia. It is important to remark that vital dynamics do not play an important role since the time scale of births and deaths is much longer than that related to SARS-CoV-2 disease progression. There are several interesting mathematical models where the vital dynamics is not taken into account due to the aforementioned facts [95, 53, 28]. We take into account vital dynamics just to be more accurate with the real situation, even though it is not a main factor. As expected, there are some uncertainties related to data of the COVID-19 pandemic which is usual in many epidemics due to, among other things, to testing surveillance [126, 123, 105]. For instance, the reported infected cases have uncertainties due to many factors such as sensitivity and specificity of COVID-19 tests [7, 126]. Moreover, the proportion of asymptomatic cases suffers from a great uncertainty [22, 36, 44, 51, 87, 96, 122, 132, 4]. Taking into account these uncertainties and using Colombia as a particular case study we set the initial conditions presented in Table 2. The total initial population *N* (0) is taken from the current Colombia population [143]. The birth and death rates are obtained from the World Bank website for Colombia [133].

**Table 2:** Initial conditions assumed for the different subpopulations using the current situation in Colombia (10th of February of 2021).

| Parameter | Symbol | Value |
| --- | --- | --- |
| Infected 1 (symptomatic) | $I1(0)$ | 35,000 |
| Infected 2 (symptomatic) | $I2(0)$ | 5 |
| Latent 1 | $E1(0)$ | 52,000 |
| Latent 2 | $E2(0)$ | 5 |
| Asymptomatic 1 | $A1(0)$ | 35,000 |
| Asymptomatic 2 | $A2(0)$ | 5 |
| Hospitalized | $H(0)$ | 2,589 |
| Recovered | $R(0)$ | 4,160,000 |
| Susceptible | $S(0)$ | 46,054,839 |
| Total population | $N(0)$ | 50,339,443 |

We use the data reported for Colombia for the 15th of February of 2021, available in different sources [59, 86]. Three key initial subpopulations are those corresponding to the infected, latent, and asymptomatic, since they affect the initial dynamics of the COVID-19 pandemic when the new SARS-CoV-2 variant is introduced. We took the seven day average of the approximated infected reported cases and then multiplied by seven days (assumed infectiousness period) to estimate the initial number of symptomatic cases of the previous SARS-CoV-2 variant. This implies that we are assuming that all the symptomatic cases are in the reported cases and the asymptomatic carriers are not reported due to low random test surveillance in Colombia [10, 2, 18, 85, 121, 16, 154, 58, 119, 37]. The percentage of asymptomatic cases in the official statistics varies for each country. In some countries it may be close to zero, since no random surveillance tests are performed. We approximated this value by relying on data from different studies and taking as the base scenario a 50% probability of being an asymptomatic carrier [10, 2, 85, 121, 16, 154, 58, 119, 37]. However, in our simulations we varied this parameter through reasonable values to take into account the uncertainty in the scientific literature regarding this percentage [11, 22, 36, 44, 51, 87, 96, 122, 132, 4]. For the initial latent subpopulation *E*1(0) we take into account that the latent period is around 5.2 days and the latent stage includes individuals who will become either asymptomatic or symptomatic [32, 78, 71, 131]. For the initial hospitalized subpopulation *H*(*t*) we rely on official data from Colombia [86]. We used another approach taking into account that hospitalized people spend an average of 10.4 days in the hospital and that around 4% of the symptomatic infected transit to the hospitalization phase and both approaches return similar results [86, 32, 78, 49]. For the initial recovered subpopulation *R*(0) we use the total of reported infected cases on February 10th of 2021 and multiply that by two in order to take into account the asymptomatic recovered cases that might not be reported in Colombia. Finally, we compute the initial susceptible subpopulation using the fact that *S*(0) = *N* (0) − *E*1(0) − *I*1(0) −*A*1(0) −*E*2(0) − *I*2(0) − *A*2(0) −*R*(0) −*H*(0). In Table 2, we present the initial conditions for the different subpopulations.

For the baseline scenario, we assume that the initial subpopulation *I*2(*t*) related to the new SARS-CoV-2 variant is just five persons. Thus, in order to be consistent with the previous paragraph we assume that there are five asymptomatic carriers and five latent individuals for the variant 2. These previous assumptions imply that the new variant VOC-202012/01 of SARS-CoV-2 has a low prevalence at the beginning of the study period. These initial conditions can be easily changed if the prevalence of the new variant is known.

## 3 Results

Here we present the numerical simulations of the mathematical model (1) under different potential scenarios related to the introduction of a more transmissible new SARS-CoV-2 variant. These simulations will allow us to analyze qualitatively the potential impact of the introduction of a new SARS-CoV-2 variant with a higher transmission rate on the dynamics of the COVID-19 pandemic. We use the parameter values of Table 1 and the baseline initial conditions for the subpopulations are given in Table 2. Several of these previous values are based on the demographics and current situation in Colombia (10 February 2021). However, these values can be adapted for another country.

We vary the transmission rates of the two SARS-CoV-2 variants in order to consider a variety of scenarios that take into account the uncertainty in the aforementioned factors. We compute the total number of infected cases for each variant, the total number of hospitalizations and the total number of deaths. These important metrics are used to understand qualitatively the effect of the new SARS-CoV-2 variant with higher transmission than the prevalent one. Moreover, the analysis of all the considered scenarios allows us to understand and observe potential situations that would need special attention from health authorities from the government and the people.

For the potential scenarios we combine variations in the contagiousness of the two SARS-CoV-2 variants and the ratio between the transmissibility of the new SARS-CoV-2 variant (VOC-202012/01) and the previously prevalent SARS-CoV-2 variant. In all the scenarios studied we vary the parameter values of the admissibilities *β*_*I*1_, *β*_*A*1_ in the range of [0.1 − 0.3] since previous studies regarding SARS-CoV-2 spread in human populations have found or used this range. In regard to the key parameter related to the contagiousness of the new SARS-CoV-2 variant we consider that is between 0% − 100% more transmissible. Thus, the ratios *β*_*I*2_*/β*_*I*1_ and *β*_*A*2_*/β*_*A*1_ vary in the range of [1.0 − 2.0]. Despite the uncertainty in these values, some researchers have mentioned that the new SARS-CoV-2 variant (VOC-202012/01) is in the range of 20%-70% more transmissible, but further research is needed to reduce the uncertainty [66]. The constructed model allows us to vary these percentages if more accurate information becomes available. Regarding the particular value of the transmission rate *β*_*I*1_ we use a range of values that have been estimated in several examples related to the COVID-19 pandemic [28, 95, 53]. It is important to remark that even though the transmission rates may not be very accurate, our approach helps to understand how a new more transmissible SARS-CoV-2 variant would affect the dynamics of the COVID-19 pandemic.

As is well known, the value of the SARS-CoV-2 virus transmission rate plays an important role in the value of the basic reproduction number ℛ_0_ [8, 47, 26, 25]. On the other hand, the effective reproduction number ℛ_*t*_ is time-varying and depends on the basic reproduction number ℛ_0_. Thus, when we consider different values of the SARS-CoV-2 virus transmission rate between humans we are implicitly considering different effective reproduction numbers ℛ_*t*_. For instance, under certain conditions ℛ_*t*_ = ℛ_0_*S*(*t*)*/N*, which relates the value of the virus transmissibility *β* to the effective reproduction number [146].

The objective here is to test the impact of the introduction of a new SARS-CoV-2 variant with higher transmissibility than the original, or previously prevalent, one on the dynamics of the SARS-CoV-2 virus. We assess this impact using the prevalence of each variant, hospitalizations, and deaths. These metrics are useful and relevant to health authorities and people in general. This approach or methodology can be extrapolated to other settings.

### 3.1 Numerical simulation of scenarios

Here we present the results of the numerical simulations for different scenarios varying the contagiousness of the two SARS-CoV-2 variants and the ratio between the transmissibility of the new SARS-CoV-2 variant (VOC-202012/01) and the previously prevalent SARS-CoV-2 variant. We obtained the total infected people, infected (symptomatic plus asymptomatic) people with variants 1 and 2, hospitalized, recovered and deaths, for all the different scenarios. Here we show the numerical results for the most important metrics and for the more interesting cases. Nevertheless, we show all the results using two parameter-space graphs for all the aforementioned populations. All the results are obtained using a study period of one year.

Table 3 shows the total number of infected (symptomatic plus asymptomatic) people for the different transmission rates of the two SARS-CoV-2 variants. It can be seen that the impact of the transmission rate is large as is usual in epidemic models, since the effective reproduction number ℛ_*t*_ strongly depends on the transmission rate. For instance, observing the first row of Table 3 it is seen that the total number of infected people is 1,271,551 under a scenario with a transmission rate of *β*_*I*1_ = 0.15 and a transmissibility rate of the new variant being the same (*p* = 1.0) as the previous prevalent variant. Additionally, it is seen that this total becomes 4,608,728 if the transmission rate is *β*_*I*1_ = 0.17 (*≈* 13% increase), and assuming same transmissibility of both variants. On the other hand, the total number of infected people is 1,879,443, under a scenario with a transmission rate of *β*_*I*1_ = 0.15 and a transmissibility rate of the new variant being 25% (*p* = 1.25) greater the previous prevalent SARS-CoV-2 variant. The variation is greater when the transmission rate of all the variants increases, which is expected. Thus, it can be seen that the introduction of a new SARS-CoV-2 variant that is more transmissible would increase the total infected population. It is important to remark that these results are obtained by the introduction of a very small number of infected people (5 infected symptomatic, 5) with the new SARS-CoV-2 variant. Table 3 also shows that when the transmission rate is small the effect of the transmission of the new variant is larger. On the other hand, when the transmission rate is large the effect of the transmission of the new variant is smaller, but still significant (*≈* 50%). These results are due to the fact that for high transmissible scenarios the number of susceptible people decreases much faster, and then the fuel of the pandemic is reduced by the nonlinear term of the transmission terms (*β*_*i*_*S*_*i*_*I*_*i*_). Thus, in a region where the transmission rate of SARS-CoV-2 has been high is under less risk for a new variant in comparison with one where the transmission rate of SARS-CoV-2 has been low. In some sense, this is the well-known herd immunity effect.

**Table 3:**
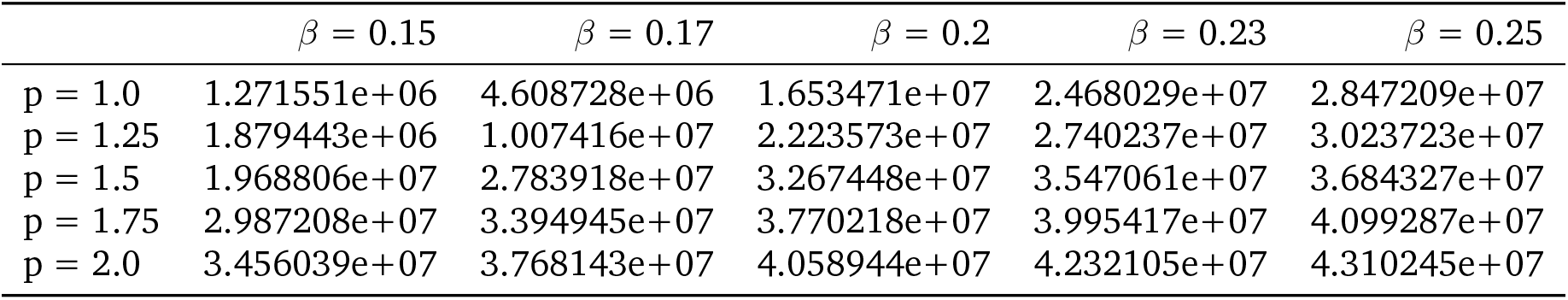
Impact of the transmissibility of the new SARS-CoV-2 variant on the total infected people (I1 + I2 + A1 + A2). In these scenarios, the transmissibility of the new SARS-CoV variant is given by p β, where β = β_I1_ = β_A1_ is the transmissibility of the previous prevalent variant. Here, we consider that the percentage of infections that are asymptomatic is 50%.

Table 4 shows an important and interesting case since the total number of infected (symptomatic plus asymptomatic) people with the new SARS-CoV-2 variant can be seen. In general when the transmission rate and the transmissibility of the new variant is increased the total of infected people with the new SARS-CoV-2 variant increases. However, for *p* values of 1.25, 1.5 and 1.75 an increase in the transmission rate is not always associated with an increase in the number of infected (symptomatic plus asymptomatic) people with the new SARS-CoV-2 variant. This result is very interesting since it means that the relationship between the transmission rates and the infected people with the new SARS-CoV-2 variant is not strictly monotone. In other words, there are combinations of transmission rates for the new and previous SARS-CoV-2 variant that make the number of people infected with the new SARS-CoV-2 variant not to increase when the transmission rate increases. The explanation for this is that when the transmission rates are large, the number of susceptible individuals decreases faster, and then there is only a small amount of people that can be infected when those infected with variant 2 is comparable to those infected with variant 1. However, if the transmission rates are very large and the second variant is very contagious then the number of people infected with this variant increases very fast at the beginning when there is a great amount of susceptible individuals. The number of susceptible individuals is the fuel for the epidemic: without susceptible individuals the epidemic cannot grow, even with a high transmission rate. This, in some way, is what happens with the herd immunity effect [47, 17, 97, 65]. However, this is not necessarily good news from a public health point of view, since governments and people are interested in the total infected people regardless of which variant the people do have. Thus, when a new SARS-CoV-2 variant is introduced and this variant is more contagious then the total of infected people would increase. Moreover, the number of hospitalized and deaths would increase unless the new SARS-CoV-2 variant causes less severity and is less deadly.

**Table 4:** Impact of the transmissibility of the new SARS-CoV-2 variant on the total infected people with the new SARS-CoV-2 variant (I2 + A2). In these scenarios, the transmissibility of the new SARS-CoV variant is given by β_I2_ = p β, where β = β_I1_ is the transmissibility of the previous prevalent variant.

| | $\beta = 0.15$ | $\beta = 0.17$ | $\beta = 0.2$ | $\beta = 0.23$ | $\beta = 0.25$ |
| --- | --- | --- | --- | --- | --- |
| $p = 1.0$ | 1.531743e+04 | 5.600301e+04 | 2.024257e+05 | 3.038535e+05 | 3.516916e+05 |
| $p = 1.25$ | 6.311486e+05 | 5.972911e+06 | 9.397759e+06 | 6.610649e+06 | 5.402739e+06 |
| $p = 1.5$ | 1.853126e+07 | 2.515835e+07 | 2.581866e+07 | 2.330687e+07 | 2.117589e+07 |
| $p = 1.75$ | 2.885323e+07 | 3.199643e+07 | 3.349876e+07 | 3.281885e+07 | 3.175012e+07 |
| $p = 2.0$ | 3.365184e+07 | 3.612819e+07 | 3.763003e+07 | 3.758772e+07 | 3.708030e+07 |

Finally, Table 5 and Table 6, show the number of hospitalized individuals for the whole study period (one year) and the number of deaths respectively. As it has been mentioned before and expected, when a new SARS-CoV-2 variant is introduced and this variant is more contagious then the total of hospitalized individuals and deaths increases, under the assumption that the new variant causes the same severity as the previous prevalent SARS-CoV-2 variant.

**Table 5:** Impact of the transmissibility of the new SARS-CoV-2 variant on the total hospitalized people. In these scenarios, the transmissibility of the new SARS-CoV variant is given by β_I2_ = p β„ where β = β_I1_ is the transmissibility of the previous prevalent variant.

| | $\beta = 0.15$ | $\beta = 0.17$ | $\beta = 0.2$ | $\beta = 0.23$ | $\beta = 0.25$ |
| --- | --- | --- | --- | --- | --- |
| $p = 1.0$ | 4.957405e+04 | 1.711537e+05 | 6.127556e+05 | 9.164529e+05 | 1.057023e+06 |
| $p = 1.25$ | 7.065646e+04 | 3.612623e+05 | 8.224247e+05 | 1.017144e+06 | 1.122371e+06 |
| $p = 1.5$ | 7.127291e+05 | 1.031755e+06 | 1.212608e+06 | 1.316221e+06 | 1.367059e+06 |
| $p = 1.75$ | 1.108271e+06 | 1.259865e+06 | 1.398868e+06 | 1.482267e+06 | 1.520734e+06 |
| $p = 2.0$ | 1.282502e+06 | 1.398100e+06 | 1.505794e+06 | 1.569921e+06 | 1.598859e+06 |

**Table 6:** Impact of the transmissibility of the new SARS-CoV-2 variant on the total number of deaths. In these scenarios, the transmissibility of the new SARS-CoV variant is given by β_I2_ = p β, where β = β_I1_ is the transmissibility of the previous prevalent variant.

| | $\beta = 0.15$ | $\beta = 0.17$ | $\beta = 0.2$ | $\beta = 0.23$ | $\beta = 0.25$ |
| --- | --- | --- | --- | --- | --- |
| $p = 1.0$ | 5345.659907 | 17520.293341 | 62924.410368 | 94605.190134 | 109116.091892 |
| $p = 1.25$ | 7301.442961 | 35135.420382 | 84220.953642 | 104951.330011 | 115841.614890 |
| $p = 1.5$ | 69942.596753 | 106053.317619 | 125125.617692 | 135814.208170 | 141050.970076 |
| $p = 1.75$ | 114227.805690 | 130003.038329 | 144326.962036 | 152915.947908 | 156877.382131 |
| $p = 2.0$ | 132337.901616 | 144247.831334 | 155338.785818 | 161942.866232 | 164922.963036 |

Finally, we show the qualitative results in a graphic form for different scenarios varying the contagiousness of the two SARS-CoV-2 variants and the ratio between the transmissibility of the new SARS-CoV-2 variant (VOC-202012/01) and the previously prevalent SARS-CoV-2 variant. Figure 2 shows the different outcomes for a wide range of different transmissibility rates for both variants. When *p* = 1 both variants have the same transmissibility.

**Figure 2:**
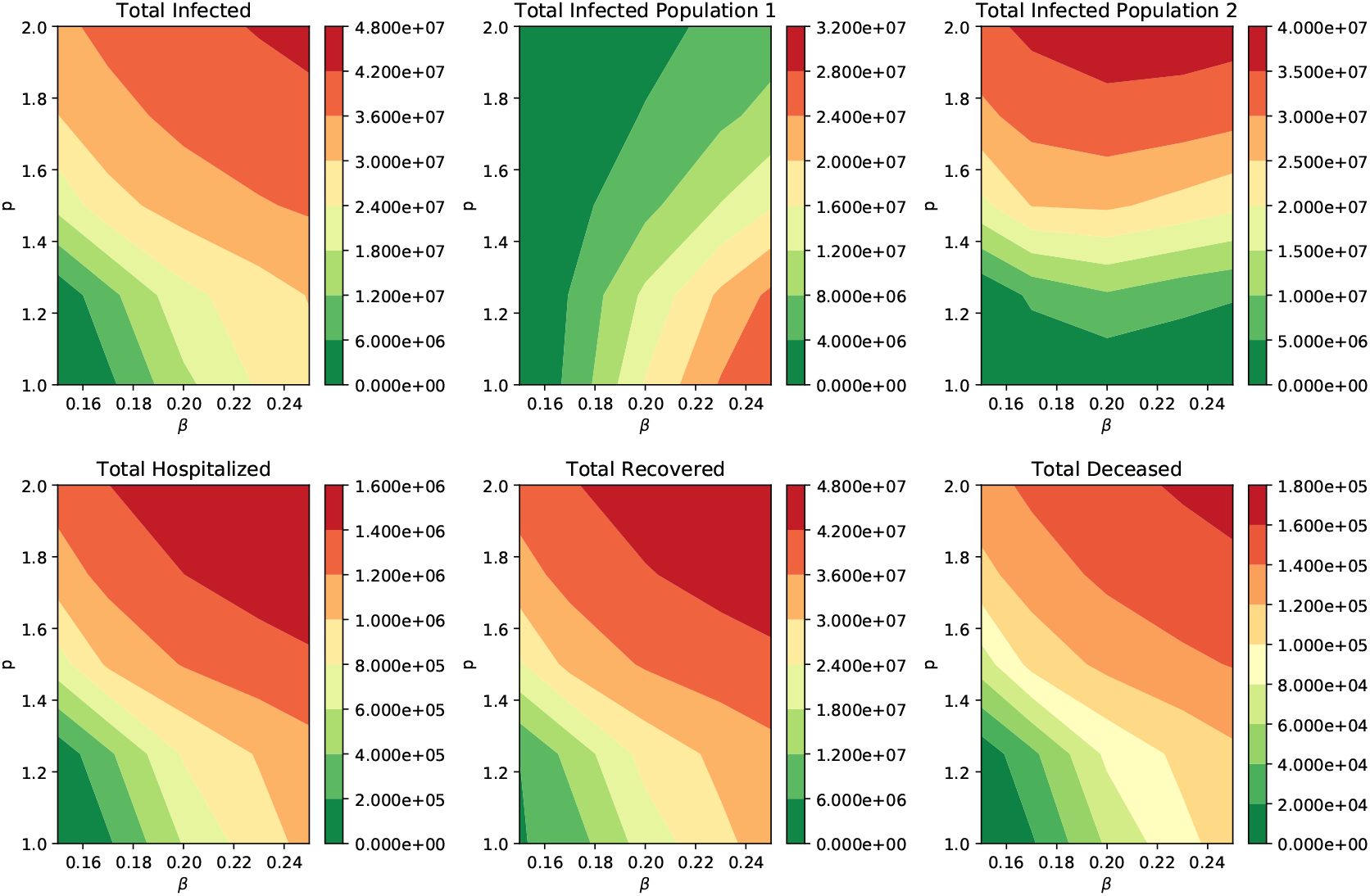
Impact of the transmissibility of the new SARS-CoV-2 variant on the total infected people (I1 + I2 + A1 + A2), total infected people with the previous prevalent SARS-CoV-2 variant (I1 + A1), total infected people with the new SARS-CoV-2 variant (I2 + A2) and hospitalized subpopulations. In addition, on the deaths and recovered cases.

As the previous Tables show, when the new SARS-CoV-2 variant is introduced and this variant is more contagious, then the total of infected people increases. Moreover, the number of hospitalized, recovered, and deaths increases.

However, for *p* values of 1.25, 1.5 and 1.75, when the transmission rate is increased, the number of infected (symptomatic plus asymptomatic) people with the new SARS-CoV-2 variant does not always increase. There is a regime where the solution is not monotone as some would expect. This behavior is not easy to grasp without the help of a mathematical model and computational tools. Here we can see one of the main advantages of the mathematical and computational approaches used in this study.

We performed many additional numerical simulations varying the parameters and the qualitative results stayed invariant. However, we added one more parameter-space graph to see the qualitative behavior after 100 days. This scenario can be seen in Figure 3. For low transmissibility the number of infected people with the new SARS-CoV-2 variant is lower than the infected people with the preexistent variant. This result is expected since the introduction of the new variant comes from few individuals (5 infected, 5 asymptomatic) and it takes time for the new variant to overcome the preexistent SARS-CoV-2 variant. On the other hand, if the transmissibility of SARS-CoV-2 is high and the transmissibility of the new variant is greater than that of the preexistent one, then the new variant will overcome the prevalence of the preexistent variant. In this regime the number of the infected people with the new variant shows a monotone behavior. As can also be seen, the total hospitalized individuals and deaths over the 100 days increase as the transmissibility of the new SARS-CoV-2 increases. This shows the risks of the occurrence of highly transmissible new SARS-CoV-2 variants. Moreover, in just 100 days the prevalence of the new SARS-CoV-2 variant overpasses the preexistent variant with a low value of the transmission rate of *β*_*I*1_ = 0.12 and assuming that the new variant is 70% more transmissible.

**Figure 3:**
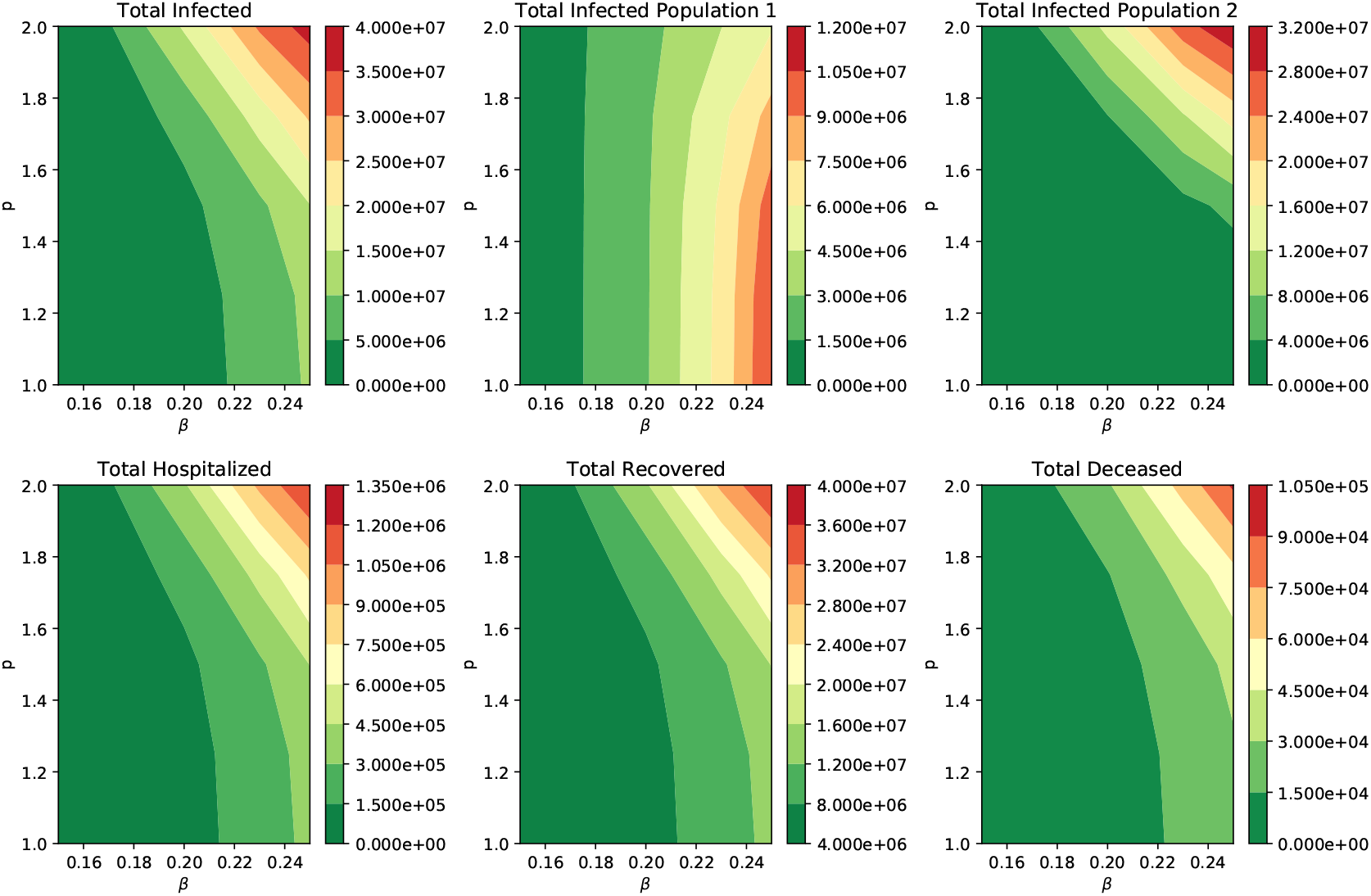
Impact of the transmissibility of the new SARS-CoV-2 variant on the total infected people (I1 + I2 + A1 + A2), total infected people with the previous prevalent SARS-CoV-2 variant (I1 + A1), total infected people with the new SARS-CoV-2 variant (I2 + A2) and hospitalized subpopulations. In addition, on the deaths and recovered cases.

## 4 Discussion

Based on the previous results we can conclude that under some plausible scenarios that the impact of the introduction of a new SARS-CoV-2 variant that is more transmissible than the previous variant would cause a larger burden of the COVID-19 pandemic. These results suggest that health authorities should focus on increasing the degree or intensity of non-pharmaceutical interventions in order to avoid more infected people, hospitalizations, and deaths. The results show that even if the new SARS-CoV-2 variant is not deadlier than the previous one, there would be more deaths. This answers questions raised recently by the media and people about the effect of the circulation of the new SARS-CoV-2 variant VOC-202012/01 of lineage B.1.1.7. As has been mentioned in the introduction some simple real-world processes are not easy to understand without mathematical models and computational tools [1]. Thus, in this article we rely on a mathematical model and numerical simulations to provide scientific support to health policies related to the COVID-19 pandemic. In particular, in this study, we constructed a mathematical model to study the impact of the introduction of a new SARS-CoV-2 variant. We used important health metrics such as total infected people, hospitalization, and deaths, in order to provide qualitative results and bring awareness of potential critical situations in many countries where the vaccination programs have not started or are in very early stages.

An optimal vaccination program helps to reduce the transmission of the SARS-CoV-2 virus in the population in an efficient way [6, 21, 64, 67, 136, 145]. The mathematical model proposed here does not consider a vaccination program since some countries have not started it yet or the number of vaccinated people is very low. We expect to construct a mathematical model in which vaccination programs are considered. The approach would be challenging due to the fact that different countries and regions have different vaccination programs, and moreover the inoculation rates do not have good consistency.

In this article, we considered the particular scenario of Colombia, but the methodology and approach presented here can be extrapolated to other countries or regions. We were able to study different potential scenarios regarding the burden of the COVID-19 pandemic. We varied the SARS-CoV-2 virus transmission rates with two different variants. We found that the introduction of a new SARS-CoV-2 variant has a high impact on the outcome. In particular, it would increase the number of infected people, hospitalized, and deaths. Another important finding is that the new SARS-CoV-2 variant that is more transmissible might become more prevalent in the population, even assuming that is 20% more transmissible. For larger values of the transmission rate of the previous SARS-CoV-2 this overtaking process would take longer since we assume that few people have the new variant at the beginning. This could happen for instance when people carrying the SARS-CoV-2 new variant travel from the United Kingdom to other countries like Colombia. This result might be counterintuitive, and by the proposed mathematical and computational approach we are able to characterize this dynamic.

The constructed compartmental model is a *SEIAR* type but with some additional features such as the compartment for asymptomatic cases. The SARS-CoV-2 virus spread is mainly driven by the values of the parameters, which have some uncertainty, as is usual in this type of model. The parameter values were chosen from scientific literature. Despite the limitations of this type of mathematical model, they have been useful in many epidemics and are a classical method to deal with epidemics [47, 15, 20, 38, 26, 9, 27, 68, 8]. Some particular limitations of this type of study are the assumption of homogeneous mixing, exponential transition times, and time-invariant parameters. Additionally, the behavior of individuals is averaged in order to avoid more complex models that in turn have their own limitations. For instance, individual agent based models have many parameters and in several cases the values of these parameters are difficult to obtain and identifiability issues usually are generated [109, 116, 39]. Despite the classical limitations of our type of model, we found valuable qualitative results that are still valid under changes of some parameters and initial conditions. These qualitative results bring awareness of the potentially dangerous situation that could arise in several countries if a new more transmissible variant such the VOC-202012/01 of lineage B.1.1.7. is introduced in the population.

It has been found that the new variant VOC-202012/01 of lineage B.1.1.7. is more transmissible [93, 75, 140, 19]. Furthermore, some researchers have predicted that this new variant will likely become the dominant variant in many U.S. states by March, 2021, leading to further surges of COVID-19 in the country, unless urgent mitigation efforts are immediately implemented [140]. The results presented here agree with this statement under the scenario that no vaccination programs are implemented or if the vaccination rate is very weak. In addition, some very recent studies suggest that the new variant VOC-202012/01 of lineage B.1.1.7. might be more deadly [52, 103, 55]. However, due to the current amount of available data further studies are needed to corroborate this [54]. For instance, some studies did not find an increase in the case fatality rate for the new variant VOC-202012/01 [14]. Another important finding is that in just 100 days the prevalence of the new SARS-CoV-2 variant overpasses the preexistent variant with a low value of the transmission rate of *β*_*I*1_ = 0.12 and assuming that the new variant is 70% more transmissible. In addition, we the results of this study show that in a region where the transmission rate of SARS-CoV-2 has been high is under less risk for a new variant in comparison with one where the transmission rate of SARS-CoV-2 has been low. In some sense, this is the well-known herd immunity effect [47, 9, 17, 65, 97]

New variants are being detected and more are expected to arise due to natural evolution [102]. In fact, the New and Emerging Respiratory Virus Threats Advisory Group (NERVTAG) has named one additional SARS-CoV-2 variant under investigation and one additional variant of concern [103]. This recently detected variant is the VUI202102/01. It is derived from lineage A.23, which is seen internationally, but the E484K additional mutation on this lineage has only been seen within the UK [103]. It was first identified by Public Health England (PHE) on 10 January 2021, while investigating a cluster of 5 cases linked to members of staff from a hospital in Liverpool. VOC202102/02 is a specific cluster characterised by the presence of the E484K spike protein mutation on the VOC202012/01 SARS-CoV-2 B1.1.7 variant that was first detected in the UK at the end of 2020.

Despite more than 4,000 SARS-CoV-2 variants having been identified across the globe, our study is valuable in various ways. For instance, even if a region has several variants circulating in the population, but all these variants have similar transmissibility (and severity) and a new variant is more transmissible like VOC-202012/01, then this study is applicable. Moreover, the results presented in this study can be generalized for regions with SARS-CoV-2 variants with different transmission rates.

Finally, our study encourages governments and their health institutions to increase the pace of the vaccination in the population in order to decrease the likelihood of a more catastrophic COVID-19 pandemic due to the occurrence of more transmissible SARS-CoV-2 variants [75]. Although we cannot predict new SARS-CoV-2 variants that are more transmissible and more deadly, this type of study brings insight into the understanding of potential future scenarios regarding the COVID-19 pandemic. Moreover, there are caveats regarding the possible protection of vaccines against new variants, and this can be studied with different mathematical models and computational tools.

## Data Availability

Data is public from different sources.

